# Protocol: Trust Your Gut: An Analysis of Dermatologic Diagnostic Accuracy

**DOI:** 10.1101/2024.03.27.24304982

**Authors:** Dana Jolley, Abraham M. Korman

## Abstract

**Background:** The current clinical misdiagnosis rate among all medical specialties is approximately 10-15%, but diagnostic error within the field of dermatology has not been studied thoroughly^1,2^. As a field that relies heavily on visual perception, many physicians consider clinical intuition to be advantageous in diagnosing skin diseases and consider it to be a rapid and unconscious phenomenon^7^. Therefore, too much contemplation may lead to more incorrect diagnoses^4^. However, while clinical intuition is a valuable clinical tool, it is widely considered to be developed throughout medical training and only successfully employed by experienced attending physicians, perhaps due to experiential knowledge and associated confidence^1,2,5^. One may expect that self-reported confidence in diagnosis would correlate with diagnostic accuracy, but this is not supported in the literature^9^. The focus of our study is to examine the development and reliability of clinical intuition as well as associated self-reported confidence levels in diagnoses at different levels of medical training among dermatologists.

**Methods:** Approximately 20 dermatologists who are PGY-2 or higher will be recruited for study participation via email. Participants will be sent a Qualtrics survey at two separate time points with a month waiting period in between. The survey will contain demographics questions, photos of 10 different dermatologic conditions for dermatologists to diagnose, and a self-reported confidence level for each diagnosis. The first survey will allow 5 seconds to evaluate a clinical photo prior to diagnosis, and this timeframe will be extended to 15 seconds in the second survey. The second survey will contain the same diagnoses, but with different pictures to avoid recall of specific photos. Following completion of all surveys, descriptive statistics will be completed with goal of publication.

**Discussion:** This study has the potential to provide invaluable information regarding the development of clinical intuition among dermatologic physicians while also examining their confidence levels and likelihood of changing correct diagnoses when given more time to ruminate. It is possible that physicians are more likely to second guess original diagnoses based off of certain demographic factors, as one systematic review found that women in medicine perceive their clinical performance as deficient more often than men^10^. Therefore, this study may give insight to the ways that complicated societal factors contribute to clinical decision making. Data from this study may be used to aid dermatologists in understanding their thought processes when diagnosing patients, and may be useful in developing education curriculum. The protocol will hopefully serve as a blueprint for creation of studies in a multitude of fields, ultimately leading to better understanding of clinical decision making and, thus, improved patient care.

## Introduction and Rationale

The current clinical misdiagnosis rate among all medical specialties is approximately 10-15%^1,2^. Diagnostic error leads to a delay in treatment for patients, unnecessary medical care, and increased health care spending. One article published on cellulitis even suggested that misdiagnosis led to approximately 50,000-130,000 unnecessary hospitalizations and 195-515 million dollars in health care spending that could have been avoided^3^. Within the field of dermatology, diagnostic error is a serious issue that has not been studied thoroughly^1,2^. In order to determine the root cause of dermatologic misdiagnosis, it is prudent to examine the process of diagnosing skin disease at all levels of training and to consider social factors that could potentially lead to medical error.

Previous studies have shown that deliberate reflection on initial diagnosis increased diagnostic accuracy within the fields of internal medicine and general practice, but fields like dermatology that rely heavily on visual perception, mental schemas, and pattern recognition for diagnosis, too much contemplation may lead to more incorrect diagnoses^4^. However, some studies assert that dermatologic diagnoses do require a great deal of deliberation and discourage diagnostic shortcuts in order to avoid diagnostic errors^1,2,5^. Others describe the visual recognition involved in dermatology as an instantaneous process^6^. In fact, many physicians themselves consider clinical intuition to be advantageous in diagnosing skin diseases and consider it to be a rapid and unconscious phenomenon^7^. Given these two differing processes for determining a dermatologic diagnosis, it is important to explore the development of clinical intuition.

While clinical intuition is a valuable clinical tool, it is widely considered to be developed throughout medical training and only successfully employed by experienced attending physicians^1,2,5^. Most medical students and residents cannot rely on intuition or gut feelings due to a lack of experience and retrospective analysis of cases^8^. This assertion is supported by cognitive science research into the diagnostic process which emphasizes that experiential knowledge is committed to long term memory and then accessed for classifying future diagnoses^5^.

Progression through medical training and development of additional diagnostic skills can lead to an expectation of increased confidence in one’s medical capabilities. This raises the additional question of whether self-reported confidence in diagnosis has any correlation with decreased medical error. Surprisingly, a previous study found that a physician’s level of confidence actually had little association with diagnostic accuracy and showed that although there was wide variation in clinical diagnosis, there was very little difference in self-reported confidence^9^. Although confidence does not seem to give way to more accurate diagnoses, it could potentially interfere with trusting one’s own intuition. This may cause physicians from underrepresented groups to second guess their diagnoses more frequently. One systematic review found that women in medicine perceive their clinical performance as deficient more often than men, including rating themselves lower in areas of knowledge, experience, and confidence even though evidence suggests no difference in actual performance between men and women at any level of training^10^. This variation in self-reported perception of skills may lead to a higher tendency among females to change diagnoses when given more time to evaluate the medical problem at hand.

The focus of our study is to examine the development and reliability of clinical intuition at different levels of medical training by surveying resident and attending dermatologists at two separate time points to calculate accuracy of diagnoses and how frequently diagnoses are changed with increasing amounts of time. We will also collect self-rated confidence levels in diagnoses and demographic factors. We hypothesize that resident dermatologists who are early in their training will have more accurate diagnoses when given increased time to evaluate a presentation, indicating poor clinical intuition. We also suspect that more seasoned residents and attending dermatologists will have better clinical intuition as shown by more correct diagnoses at a shorter time period of evaluation. However, we expect some of these more experienced physicians to have a high likelihood to change their initial diagnosis to an incorrect one with more time to evaluate a presentation. The likelihood of second-guessing an initial diagnosis will likely be dependent on confidence or certain social factors, such as race or gender.

## Materials and Methods

### Aims

#### Primary aim

1. Determine the accuracy of resident and attending dermatologists’ clinical diagnoses versus examination time variations
2. Assess the development of clinical intuition among resident and attending dermatologists

#### Secondary aims

1. Describe the likelihood of incorrectly changing diagnoses with greater amounts of time to second guess
2. Determine the association between confidence in diagnoses and accuracy of diagnoses

### IRB Approval

This study was determined exempt from IRB review on 11/07/2023 by Ohio State University’s Office of Responsible Research Practices in qualifying exempt category 2b. Study number 2023E0952.

### Procedures

#### A. Targeted Audience

Our research is targeted to dermatologists.

#### B. Data Distribution

The data collected from this study will be used to evaluate the development of clinical intuition and examine social factors that may play into trusting that intuition. The data will be published in a dermatology journal to distribute information to practicing dermatologists.

#### C. Research Design

1. *Sites*
  a. The Ohio State University
2. *Inclusion criteria*
  a. Dermatologists who are PGY-2 or higher in level of medical training.
3. *Methods*
  a. We are designing a survey-based, crossover study in which we query approximately 20 practicing dermatologists at two separate time points.
  b. We will send a recruitment email to the dermatologists to gauge interest in participating in the study. We will include approximately 20 dermatologists who agree to participate. Recruitment will begin on 2/11/24 and end on 3/20/24.
  c. The participating dermatologists will be sent a Qualtrics survey at two separate time points. They will be sent with a month waiting period in between.
  d. The email containing the survey link will also include a copy of IRB-approved informed consent document, containing verbiage that clicking the survey link serves as consent to participate in the study.
  e. The survey will be built as follows:
    i. We will provide photos of 10 different dermatologic conditions for dermatologists to diagnose.
    ii. For the first survey, dermatologists will be given 5 seconds to evaluate the photo prior to making a diagnosis. The time will be extended to 15 seconds for the second survey.
    iii. Each survey will also gather the physician’s confidence level in each diagnosis on a scale of 1-5, with 5 being highest level of confidence.
    iv. Each survey will collect demographic information such as level of medical training, race, age, and sex.
    v. Each survey will be scored as correct or incorrect for accuracy of diagnoses using a master key.
    vi. The survey at the second time point will include the same diagnoses, but with different pictures to avoid participants recalling specific photos.
    vii. The second survey will be administered 1 month following the administration of the first survey.
    viii. Each survey will take approximately 5-10 minutes to complete.
  f. Only participants who complete the first survey will be sent the second survey. The Qualtrics survey will include their name and email, which is how we will know where to send follow up surveys.
  g. Once the surveys are completed by all dermatologists, the data will be analyzed and interpreted with goal of publication.
4. *Variables*
  a. Independent: survey questions along with a mixture of inpatient and outpatient dermatologic diagnoses
  b. Dependent: survey answers

#### D. Sample

We plan to query approximately 20 dermatologists from The Ohio State Department of Dermatology.

#### E. Measurement/Instrumentation

Descriptive statistics will be performed on results of survey questions and diagnoses.

#### F. Detailed Study Procedures

After IRB exemption, we will begin contacting dermatologists to participate in our study. All data will be viewed at the OSU facilities under a locked door and saved within the secured access dermatology research drives. Recruitment will begin on 2/5/24 and is expected to continue for approximately two months. Survey data will be analyzed once all participants complete the second survey and results will be reported via academic journal in order to disseminate information to practicing dermatologists.

#### G. Data Analysis

Primary Outcome Measures: Quantitative assessment of correct diagnoses by dermatologists at different time points.

Secondary Outcome Measures: Quantitative assessment of factors such as demographics and level of medical training which may affect one’s trust in a clinical diagnosis.

## Discussion

### Limitations

#### A. Internal Validity

Dermatologists may possess varying levels of experience with treating certain clinical diagnoses (e.g. inpatient vs outpatient). To address this internal issue, we will ensure equal numbers of diagnostic categories and recruit dermatologists from all backgrounds.

#### B. Loss to Follow-Up

Completing surveys at two separate time points may be a difficult task for many busy practicing physicians. To address this issue, we have designed the survey to be as short and user-friendly as possible. However, we still expect to lose some participants to follow-up and will try to recruit more participants than our sample size to offset this issue.

## Data Availability

No datasets were generated or analysed during the current study. All relevant data from this study will be made available upon study completion.

## Authors’ Contributions

Dana Jolley, BS; Conceptualization, data curation, formal analysis, methodology, project administration, validation, manuscript preparation

Dr. Abraham Korman, MD; Conceptualization, methodology, writing (review & editing)

